# Building contextually-relevant training programs for scientific development: process and lessons learned in implementing two iterations of a faculty enrichment program in applied malaria modeling

**DOI:** 10.1101/2025.05.20.25327477

**Authors:** Letitia Onyango, Ghislaine Ouédraogo-Ametchie, Anne Stahlfeld, Tobias M. Holden, Ricky Richter, Manuela Runge, Kok Ben Toh, Isaiah Agorinya, Benedicta Mensah, Jaline Gerardin

## Abstract

**Background:** Mathematical modeling can be a useful approach to explore potential impact of malaria interventions and thereby inform resource prioritization decisions. However, expertise for applied modeling for public health decision-making is limited in malaria-endemic countries. A 4-month faculty enrichment program (FEP) in applied malaria modeling was implemented at Northwestern University, USA, in 2022 and 2023 with components including technical skills development, communication skills development, and domain knowledge on malaria epidemiology.

**Methods:** Two cohorts of FEP participants and instructors were interviewed at baseline, midline, and endline to understand their expectations, experiences, and challenges with the program.

**Results:** Participants valued their growth in technical expertise, research skills, and communication ability, as well as clear opportunities for knowledge transfer at their home institutions. Participants reported challenges with cross-disciplinary learning, balancing program components, and adapting to new teaching and learning styles. Instructors adapted program structures and teaching approaches to adjust to participant needs and reported strengthening of their own technical capacity.

**Conclusion:** Training programs for technical skill development must be informed by the needs and priorities of prospective participants and include continuous feedback mechanisms to respond to emerging needs. Multi-pronged approaches increase long-term program value to participants and help establish pathways for knowledge transfer.

## BACKGROUND

Data analysis and mathematical modeling are increasingly being applied to inform decision-making in public health across the globe, from COVID-19 to malaria [1–3]. Scholars with skills in mathematical modeling of infectious diseases, including malaria, are present in Sub-Saharan African universities. However, challenges persist with the extent of existing modeling capacity, limited availability of training programs for malaria modeling, and successfully bridging between academia and public health [4,5].

To address these gaps, a faculty enrichment program (FEP) in applied malaria modeling was implemented at Northwestern University in 2022 (Cohort 1) and 2023 (Cohort 2). The program was designed and led by an applied malaria modeling research team, and participants were embedded within the team for 4 months. The program targeted existing faculty at Sub-Saharan African institutions who were positioned to add new directions onto an existing research program, use their university positions to facilitate relationships with public health programs, pass on skills to their own students, and collaborate scientifically with the malaria modeling research team at Northwestern University.

The FEP was cross-disciplinary, covering development of technical (coding, modeling), scientific (epidemiology, entomology, public health), and soft (oral presentations, proposal preparation) skills. Given the cross-cultural nature of the program, both participants and program instructors encountered new dynamics related to learning and working environments. Participants were therefore simultaneously challenged in many domains with which they may not have already been familiar.

As applied modeling to inform policy is maturing as a field, and the need for sub-Saharan African countries to become self-reliant in public health analytics is more urgent than ever, the experience of this faculty enrichment program can inform those seeking to strengthen public health analytical and modeling skills in Africa and elsewhere.

## METHODS

### Program structure

The multi-component faculty enrichment program (FEP) was implemented in person at Northwestern University in Chicago, USA, for 16 weeks in 2022 (Cohort 1) and 18 weeks in 2023 (Cohort 2) (Figure 1). All participants (Table 1) had completed their PhD training in African institutions and had prior research experience, including publication records in their respective disciplines. At their respective home institutions, participants were primarily engaged in teaching. Instructors were postdocs, PhD students, or research staff at Northwestern University. In Cohort 2, instructors also included one alumnus from Cohort 1 and a researcher from Ghana with previous experience with using EMOD. Each participant was paired with one instructor, with whom they met one-on-one each week for coaching, ad-hoc support, technical questions, and review of draft documents. Each participant also met with the program director one-on-one each week. Additional help was available at end-of-week group wrap-up sessions (Cohort 1) or during office hours held by staff (Cohort 2).

**Figure 1.**
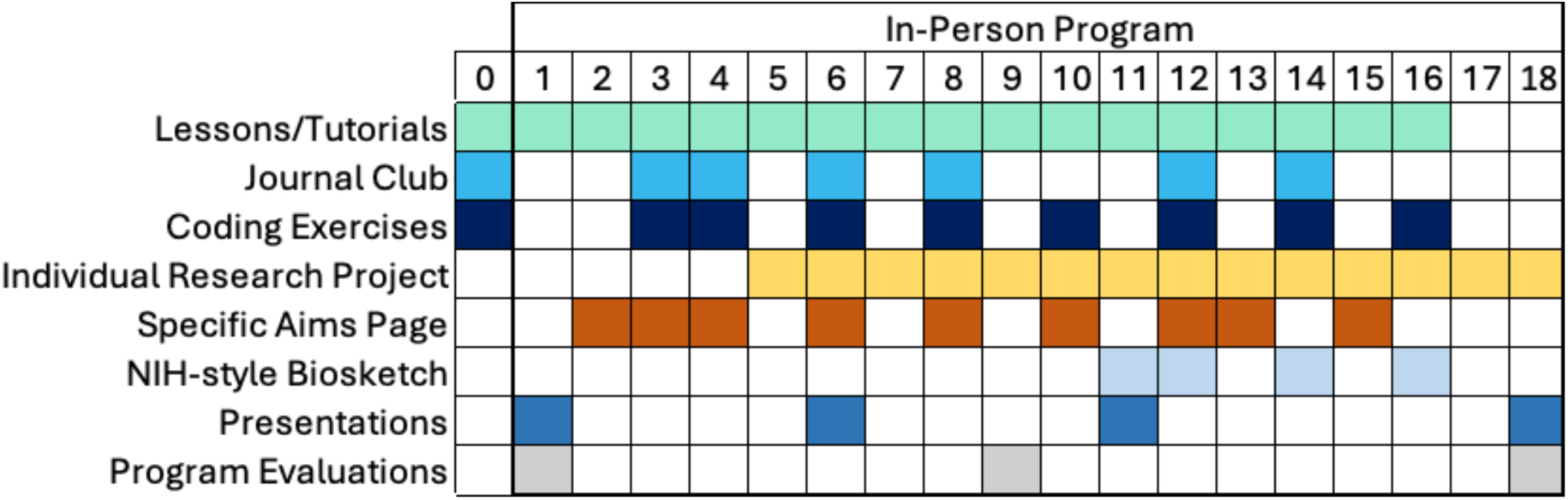
An overview of the faculty enrichment program structure, as implemented for Cohort 2.

**Table 1.**
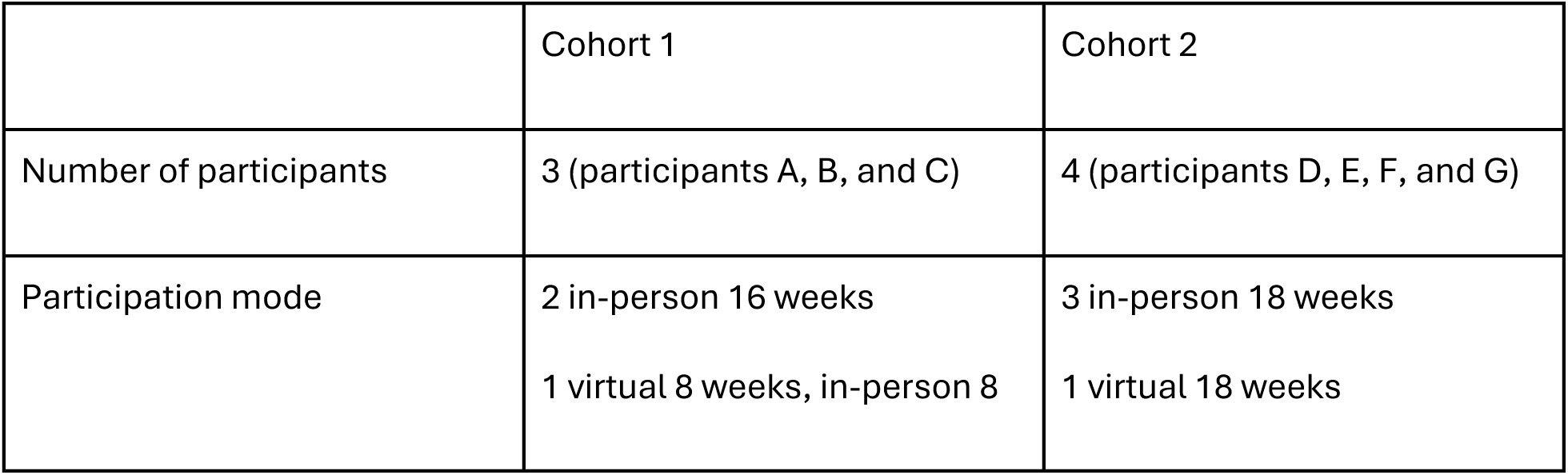
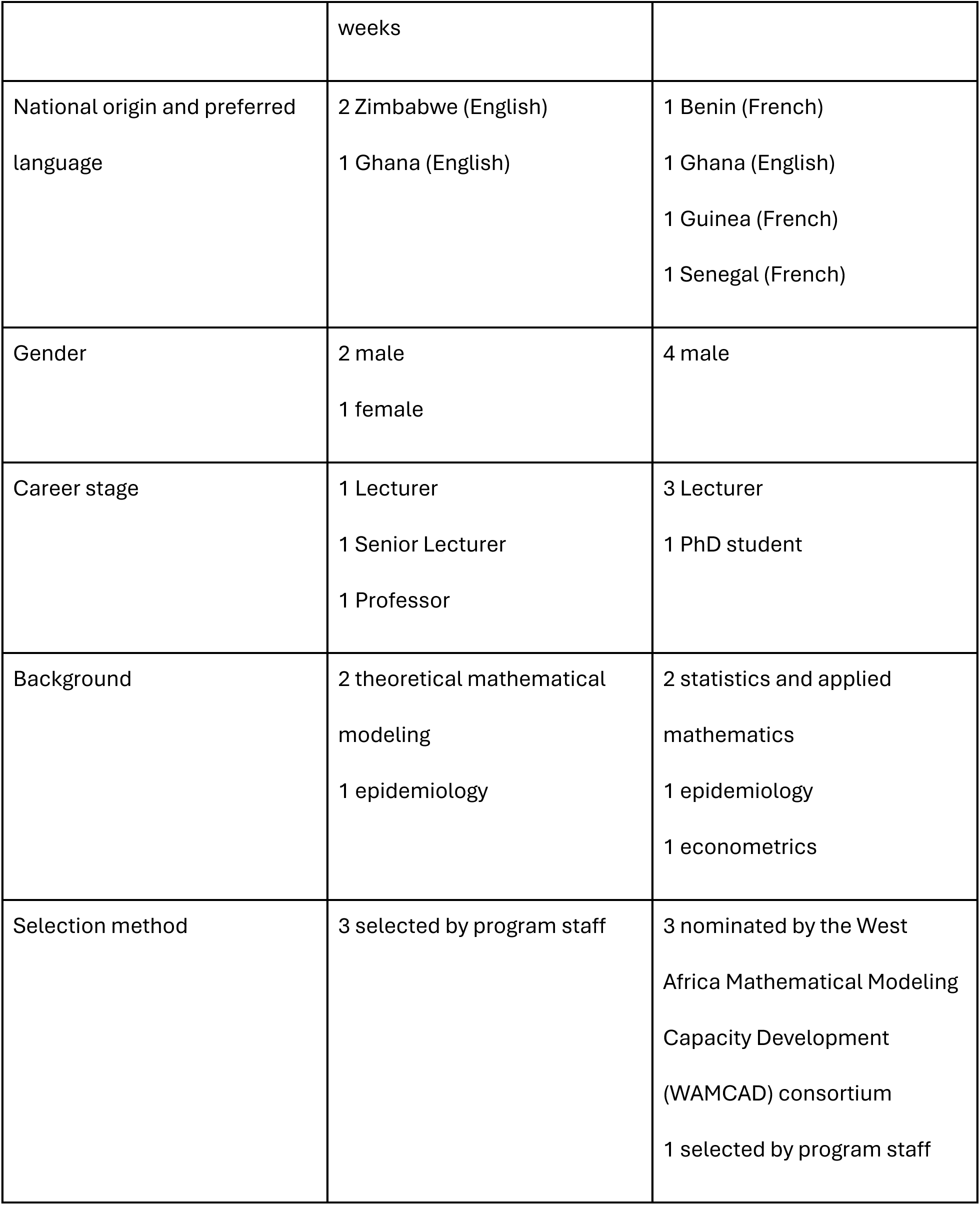
Participant characteristics for Cohorts 1 and 2.

**Technical skill development** centered around the use of the agent-based malaria model EMOD in and programming with Python. Participants learned the basics of EMOD usage through lectures and example exercises, which were conducted as a group during tutorial sessions. Each participant designed and executed a research project that included parameterizing, generating, and analysis of malaria simulations in EMOD. Supplemental lectures and coding activities covered using epidemiological survey data, map-making, and data visualization.

**Communications skills development** included preparation of a Specific Aims page, with multiple iterations of feedback and revision, based on the participant’s EMOD research project; preparation of an NIH-style biosketch (Cohort 2 only); 4 oral presentations with feedback sessions, including 3 on their EMOD research project; preparation of a poster (Cohort 2 in-person participants only); participation in feedback sessions for practice talks given by instructors on their own research; and participation in peer feedback sessions (peer editing hour) on Aims pages, biosketches, and other writing prepared by instructors on their own projects.

### Domain knowledge

All participants were new to applied malaria modeling and some were new to epidemiology. Journal clubs with an applied malaria modeling paper were held every other week, each led by a participant. Topical lectures on vector biology, within-host models, novel vector control, and other malaria models were held later in the program while participants were focused on implementing their research projects. For Cohort 2, virtual lectures on the basics of epidemiology and malaria were given the week prior to program start. Cohort 2 was also able to attend the annual meeting of the American Society for Tropical Medicine and Hygiene and a meeting of the Applied Malaria Modeling Network held in Chicago during week 17.

### Additional special sessions

Discussion sessions were held covering limitations and future of EMOD and in modelling more broadly; challenges of continuing EMOD-based research at home; and challenges specifically with high-performance computing at home. For Cohort 2, a representative from the Gates Foundation gave an overview of the foundation’s malaria priorities. Program structure was similar for Cohort 1 and Cohort 2. Changes to Cohort 2 were in response to recommendations from Cohort 1 participants and staff, and included clarification of expectations for program components, including a more extensive program handbook; a virtual week of lectures on background material; reorganization of EMOD exercises and tutorials such that they were all completed before participants began implementation of their research projects, rather than concurrently; emphasis from the beginning on use of a high-performance computing cluster; and translation of Cohort 2 written materials into French.

### Ethical review

This study was reviewed and approved by Northwestern University’s Institutional Review Board, Project STU00217422, as not constituting human subjects research. Participants provided informed consent.

### Study design

All FEP participants participated in in-depth interviews at baseline (week 1), midline (week 8) and endline (week 16 or 18). These interviews recorded participant experiences longitudinally, highlighting changes in perceptions of program content, skill development, and relationships with instructors. At baseline, participants were asked to reflect on their motivations for participation, their expectations, and their post-program plans. At midline, participants were asked to reflect on their overall impressions of FEP content and structure, challenges, self-assessments, and their expectations for the remainder of FEP. At endline, participants were asked to reflect on the extent to which their participation in FEP met their individual professional objectives and to provide recommendations for future iterations of the program. Participant interviews were supplemented by interviews with instructors at baseline, midline, and endline. Instructor interviews covered participant progress, program challenges, and how core assumptions affected program implementation. Interviews were administered by two of the authors (LO and GOA) in either English or French depending on the participant’s preference. Neither interviewer had any other role as FEP staff.

### Data analysis

Interviews were professionally transcribed and checked by members of the research team to ensure accuracy. The study team reviewed transcripts from each participant utilizing an interpretive phenomenological analysis approach [6] which allowed for participants’ subjective experiences to guide the analysis of their experiences with program structure and implementation.

## RESULTS

### Participant motivations and initial expectations

Participants were motivated by the opportunity to work on applied malaria modeling with real-world data. According to participants, data access at their home institutions was difficult, resulting in a reliance on theoretical models. In both cohorts, participants felt that the combination of training in a state-of-the-art model, access to real world data, and access to new collaborators would enable them to explore new questions about malaria in their home countries, resulting in career advancement and greater funding and publication opportunities:

> *I’m interested in moving away from just theoretical modeling into applied modeling. My expectation would be to be able to write a research proposal on the applied [modeling] and be able to attract funding for that research.* (Participant A, Baseline)

Participants in Cohort 1 added that their newfound exposure to use cases for Covid-19 modeling deepened their interest in malaria modeling and expanded their view of how modeling could support decision-making.

Prior to starting FEP, participants in both cohorts noted that while they had participated in modeling skills training programs before, the duration, depth, and approach of FEP were new to them:

> *In general, training programs for teachers are usually two- or three-day seminars. It’s very short. No time to assimilate lots of things, in general, it’s just to introduce something. If you’re not very motivated to continue yourself, it’s not going to do much for you.* (Participant F, Baseline)

> *I think the uniqueness of this program is the fact that it’s hands on. And there’s a lot of resources available for participants to explore. The community is wide, and the resources are deep.* (Participant B, Baseline)

Several participants noted that the inclusion of funding for everyday expenses was an important enabling factor for their participation, as it assuaged concerns about how they would negotiate leave from their home institutions. This was especially important for participants with family expenses to manage.

Participants expected that having a designated paired instructor with them would facilitate their success. Given the new material, it was important that they had resources to navigate challenges with their projects:

> *The fact that I’m assigned to someone who I can ask questions directly and have a chat with when I’m stranded will help me succeed.* (Participant B, Baseline)

Participants in both cohorts expressed that they did not anticipate challenges adapting to the learning system as they had prior experience collaborating with global researchers and some experience participating in trainings and workshops administered by American faculty. Among Francophone participants in Cohort 2, one participant expressed that they did not anticipate any challenges with English instruction, feeling that the provision of written training materials and supplemental resources in French would be sufficient. Two Francophone participants, however, expressed some concern about how limitations in their English-speaking capabilities might pose challenges for their success.

All participants were motivated to engage in knowledge transfer activities upon return to their home campuses. They viewed their participation in FEP as an opportunity to build the foundational skills to implement applied modeling training at home:

> *[I plan] to start teaching computational epidemiology using EMOD at my current university. Currently we aren’t running any course in mathematical epidemiology, which I feel is a gap. I think when I return to [home country] I can introduce or propose this for undergrad students.* (Participant G, Baseline)

Finally, participants expected that their involvement in FEP would enable them to work more closely with National Malaria Programs (NMPs) and be of service to their home countries. In their view, participation in FEP would give them more leverage and legitimacy when navigating conversations with the NMP. Participants noted their aspirations to collaborate with and train key NMP personnel and other ministry of health staff in the foundations of modeling so that public health practitioners could better understand the implications of modeling outputs:

> *I’m hoping I can also help train the staff of the national malaria control program. Alongside this program, there’s also the Neglected Tropical Diseases program. The interesting thing would be to organize workshops or training seminars program managers in mathematical modeling.* (Participant D, Baseline)

### Perceived value-add of FEP

#### Overall perceptions

Participants described overall positive experiences with FEP, and that their experiences far exceeded expectations. Participants cited the breadth of program activities, opportunities for collaboration with other modelers, and access to the cultural capital needed to thrive in research collaborations with global faculty. Participants appreciated the integrated curriculum that used applied malaria modeling as a framework for strengthening research skills:

> *I see it as a skill building program, in terms of research, or rather epidemiological research on malaria, specifically using mathematical modeling methods. I say this because I don’t see it as just training in mathematical modeling. I see it as a full introduction to both the design and implementation of research issues that are specific to malaria. But you could also say more generally to public health. It’s well-structured because you can’t do modeling without constructing problems. Therefore, you first learn how to identify a research problem. You learn how to build it yourself and how to identify the tools that are going to help you answer those questions. And then, you learn how to implement that. So I think it’s a complete package.* (Participant E, Endline)

#### Growth in technical expertise

All participants reported increased technical expertise in malaria epidemiology, programming skills, and EMOD use regardless of their prior experience. Participants with limited prior experience with modeling expressed growth in their modeling capabilities, such that they were able to execute their research projects:

> *I have learned a lot looking at the data and accessing the data, the true data that you need for modeling, the modeling itself, right? I have not worked with agent-based models before. Now I can interpret the results that I managed to come out with. Besides the modeling, I’ve also learnt Python programming. I have also learned R.* (Participant A, Endline)

> *The program really enabled me to reach my expectations. I had no skills in modeling whatsoever. So this program allowed me to see, really, how to work with modeling with tools I didn’t know. It really helped me achieve my aim.* (Participant D, Endline)

At midline, participants expressed a growing ability to configure and run EMOD, understand EMOD capabilities in relation to their projects, and think through their research objectives. Participants cited program structure, tutorials, and support from fellow participants as the components of FEP that improved their technical expertise:

> *The past seven weeks have been an eyeopener for me. When we started with the whole EMOD thing, it felt so massive and so enormous in structure in terms of what we are required to learn. And it was a bit confusing at the beginning trying to figure out what exactly the various components are and how they relate and how you can use them, and which ones are applicable to your project, and which are not. But going through the individual sessions, the weekly tutorials, shaped my understanding of [what] the various components are. It gave me the opportunity to be able to think about my own research topic clearly, and then outline the objectives.* (Participant B, Midline)

At endline, participants across both cohorts felt much more capable in using EMOD and expressed a desire to continue using EMOD for their research. One participant added that exposure to other EMOD inspired them to explore other models with more confidence:

> *Outside the EMOD training, we’re exposed to other models. It’s not like, hey, just come here and learn what we have. But there are other models out there that could be of interest. But I would say for someone like me, from an analytical point of view, I find EMOD more supreme because of its technicalities and how complex it is. It was just some short talks or presentations on other models and malaria simulations. [Learning EMOD first] made understanding the concept of those models much easier, because you’ve been trained on something that is a bit complex.* (Participant G, Endline)

#### Improved data access

As participants learned more about country-level data collection and potential uses for country data, they gained a greater appreciation for how they could fill gaps in data coordination and use that existed in their home countries:

> *There is this huge gap between generators of the data and users of the data. And me being here and seeing how much work – that coordination that goes around here and how the setup is, then you realize that [data coordination and use] is happening in countries where there’s no malaria. So for me, it’s not just about my career, but I think that it’s addressing a need for more experts in this area in malaria endemic countries. And the target [of FEP] is perfect because it’s targeting faculty – like training of trainers.* (Participant B, Midline)

#### Scientific development

Participants discussed how journal clubs, team meetings, and peer editing hours helped them improve their scientific communication, presentation, and critical thinking skills. Participants described how journal clubs allowed them to work together to understand the components of good scientific practices and writing and encouraged participants to think more deeply about how they communicated their own research. Participants expressed that their ability to identify effective messaging and communication in manuscripts and presentations greatly improved. The opportunity to hold in-depth discussions about papers and about the research projects of FEP instructors helped participants to develop better questions about methodology and reflect on the strengths and weaknesses of modeling projects:

> *The most fundamental thing here is the fostering of critical thinking. By reading peer-reviewed articles, by taking part in sessions where you try to review the work others are currently working on. This was very important to me and I think it sharpened my critical thinking skills.* (Participant E, Endline)

Participants reported that the inclusive nature of team meetings, peer editing hours, and practice presentations reframed their approaches to collaboration and expressed an interest in replicating these structures at home. The opportunity to engage in research discussions and feedback sessions with the entire Northwestern University team inspired participants to change their approaches to communication and developing their students:

> *Initially I had this limited way of preparing slides, but when I got here, I see the way people present their slides, how to give a talk. All these things have really reshaped my thinking in certain things, and I think it’s good. It’s something I will propagate back in my investigations.* (Participant G, Endline)

> *We are looking forward to introducing the journal club meetings and regular presentations to the group of students that we supervise.* (Participant A, Endline)

Despite some challenges with English instruction, Francophone participants also reported benefits from engaging with the scientific development components. Given that most grant proposals are in English, engaging in immersive learning and scientific review in English strengthened their language skills and subsequently, their aims pages and biosketches. One participant felt that Francophone researchers should have more access to similar opportunities to expand their access to funding opportunities:

> *In French-speaking countries, we really have a challenge. Language can pose a real difficulty for [researchers]. In the scientific world, there are a lot more grant proposals and everything in English. There’s always this language problem, I think they’re doing far more interesting things. We’re not doing everything we can so that [we can access] those grants. So the act of training modelers in languages other than English, that could be a good thing.* (Participant F, Endline)

#### Peer learning

Because participants spent the most time with other participants, they were more likely to attempt problem-solving with their peers before approaching instructors. In Cohort 1, several participants reported getting more from peer interactions than from instructors. As one instructor observed:

> *[Participant C] is less likely to take the initiative to approach [instructor] and comparatively it feels like I need to reach out to them once in a while to make sure that things are all right. So they interact more with the [participants] and learn more from that than reaching out to [me to] ask questions to help build their own project.* (Instructor B, Midline)

During Cohort 2, there were fewer mentions of peer learning during midline and endline interviews. While Francophone participants sought one another for peer collaboration, the lone Anglophone participant attributed their failed attempts at more collaboration to the other participants’ preference for speaking in French:

> *I sometimes want us to work as a team and help each other to move forward, because like my previous experience, that’s what we do. We work together, we sit together. Sometimes I’ll be like, oh, let’s meet together, let’s try to identify our issues, and let’s meet the team members during their office hours. But when I get there, no one shows up, and I only go there myself to just see them and come back and work on my project. I don’t feel it’s healthy, and I sometimes feel it’s more like a cultural difference. Since everyone has his way or objectives, I don’t want to push, because we are all adults. But it would have been nice for us to even work on the projects together.* (Participant G, Endline)

#### Knowledge transfer

At baseline, all participants were confident they would be able to lead trainings in modeling using EMOD upon returning home. At endline, participants expressed more mixed perceptions, with some noting that they needed to build on their experience before attempting to teach the full scope of EMOD principles and use:

> *I can offer the basics. I will not say I’m full - I think it’s a continuous - what I have experienced here is a bright foundation to learn more. I will not call myself a teacher in modeling right now because I still need to drop some milestones in it. I’m hoping that my collaboration with [assigned instructor] and the continuation of my project here will give me more leverage to teach. So I’m not going to rush into teaching it now. But I will try to gain relevant experience in addition. I will initiate it.* (Participant B, Endline)

Participants who were more confident in their ability to lead knowledge transfer at endline saw opportunities to replicate training modules in their labs and propose new courses focused on EMOD. These participants felt that the introduction of mathematical modeling and EMOD sessions would fill gaps in existing curricula:

> *We have colleagues who would have liked to use the technique to implement it in their research programs. So that would be the first thing, to share with all these people all the things I’ve been able to discover here, EMOD and other applications. We already have a weekly system where we share knowledge. Therefore, I’m going to integrate this training into that, where we will run a number of sessions.* (Participant E, Endline)

> *There’s one faculty in my school who is also into mathematical modeling, so currently we don’t run any mathematical modeling course, even at the postgraduate level. I’m working with him currently to develop a curriculum for something – we want to call it computational epidemiology, where we have EMOD, malaria simulation. So, it would have the first part being the normal deterministic disease modeling and EMOD would come into play at the computational aspect.* (Participant G, Endline)

### Growth in program staff

FEP spurred personal growth in instructors, including improved teaching approaches, improved capabilities in preparing teaching materials, and expanded awareness of training approaches outside the US. Several instructors noted how their own understanding of EMOD improved:

> *I realized that it was going to be a good opportunity to point out some gaps in my own understanding of things, like if I can come up with an exercise or write instructions for the how-to. So, that was helpful even before [participants] got here. [FEP] also made me realize that there are certain things that not just I can teach, but maybe I am the best person in the group to talk about this one particular part of what we do. So, some capacity building, maybe, for me as well. Or at least confidence building.* (Instructor D, Midline, Cohort 2)

Participant feedback motivated instructors to incorporate illustrative use cases for EMOD into the instructional material. Instructors could use their own research projects to illustrate each concept of EMOD being introduced and allow participants to better understand real world applications of what they were learning:

> *It would be nice to have some more use cases across the team, how EMOD is used. […] for example, this week, individual properties have been introduced, and I’m having a use case of who on the team is using this feature of EMOD… to make it less abstract, more relatable to how to use it.* (Instructor C, Midline, Cohort 1)

At the conclusion of Cohort 1, instructors were especially motivated to consider how future FEP cohorts and lab members would benefit from more robust resources and documentation.

### Challenges

Participant and instructor interviews revealed several differences in program expectations and experiences. The disciplinary background and habitual learning styles of participants had a much greater impact on their learning experiences than they or instructors had anticipated.

#### Challenges with participant preparation and cross-disciplinary learning

Disciplinary backgrounds created several challenges for participants. Across cohorts, participants and FEP instructors felt that the curriculum was sufficient for individuals with strong analytic and programming backgrounds to easily grasp EMOD and associated epidemiological concepts. However, participants with mathematical backgrounds felt that their limited knowledge of epidemiology created challenges for them and noted that a crash course would have been helpful. Instructors in Cohort 1 agreed:

> *Because neither of [two participants in cohort 1] really have epidemiology backgrounds, even though they’ve done some infectious disease work, the concept of an incidence rate is very foreign, and I have had to sit down and try to explain that. An incidence rate might be more beneficial than cases, but also sometimes cases might be more beneficial [as an outcome indicator] than an incidence rate. They’re both useful in different ways. But I think understanding what that was, and knowing where to look to understand what it was, was challenging.* (Instructor A, Endline, Cohort 1)

While an epidemiology crash course was implemented for Cohort 2, members of Cohort 2 without epidemiological backgrounds still needed additional support from instructors to improve their familiarity with concepts presented in the crash course.

Participants and instructors felt that participants would benefit from strengthening general computational skills prior to FEP start to ensure that participants could have full command of EMOD:

> *We were taught how to install EMOD, how to execute the commands. But I had a lot of trouble in the beginning because I’ve used R, EPI info, SPSS, but I had never used Linux commands. But when you run the EMOD program on Quest, you use Linux commands. So that bothered me before I was able to get the hang of it. I think that if the program could add a refresher or a review of the commands and the use of Linux commands at the start of the training, that would make it easier for a lot of people like me who have done statistics but who haven’t used Linux commands.* (Participant D, Endline)

Participants discussed how their disciplinary backgrounds had a strong impact on their approaches to learning, and how their experience of learning EMOD may have run counter to their expectations:

> *When you have someone with a biological background, especially a PhD person with a biological background, that person’s way of thinking is quite different from someone who has a mathematical background. Because mathematicians expect to see A to B to C to D. But it seems EMOD is more of like – it’s open. It’s created not to be strict in relation to the modeling approach. It’s more of like an open platform where it takes in any kind of assumption you make towards whatever you want to achieve. Before you start this program, emphasis should be placed on the fact that forget about everything you know in hardcore mathematical modeling. Someone coming to the program would think I’m coming to work with some equations and you get there, and there are no equations.* (Participant G, Endline)

Participants with a theoretical modeling background were especially surprised, as they expected to be provided with a deep review of the equations behind EMOD as part of the curriculum. However, the FEP curriculum focused on the use of EMOD rather than its source code or theoretical basis, especially during the early weeks. Later lectures covered aspects of EMOD structural assumptions, and a full reading list of EMOD papers was available to participants. However, participants in both cohorts experienced various challenges with the approach of learning model application before theory. Despite a stated desire to understand the equations behind EMOD, participants did not proactively seek out EMOD’s foundational papers or investigate the source code on their own.

#### Challenges with program components

Participants in both cohorts cited challenges in keeping up with the pace of FEP, especially where its structure required managing technical and scientific development components simultaneously. At midline, participants in both cohorts felt the rigor and pace of FEP was more difficult than they expected:

> *You end up losing focus because while you are while you are working on your project, you have another assignment, or you have another target to achieve. So it seems like you are doing a lot of things within a week, within two weeks.* (Participant G, Midline)

At midline, participants felt that scientific development components were important career boosters. However, as they started focusing more on their EMOD-based projects, the time commitment required for scientific development components felt increasingly challenging as the pressure to complete their project intensified.

#### Challenges with learning and teaching styles

Instructors in Cohort 1 assumed that participants would be proactive about seeking support outside the classroom for completing EMOD exercises and implementing projects, but participants were less proactive than expected. Instructors became anxious that participants were spending too much time slowly troubleshooting problems themselves, and therefore not progressing as rapidly as expected. Cohort 2 instructors were more aware of this dynamic and made an effort to remind participants of the availability of instructor support throughout FEP.

Participants themselves felt that they made adequate use of instructor availability when it was needed. Participants expressed a preference for sitting with the material themselves before seeking clarification for what they did not understand, rather than immediately asking questions during lectures and tutorials:

> *I think that the collaboration is very friendly and open. I’m very encouraged to ask questions. But I’m the kind of person who doesn’t ask a lot of questions. If you try to explain something to me one time, I’m trying to deeply understand it. If I understand it, good. If I don’t understand it, I can come to you.* (Participant D, Midline)

During the first few weeks, instructors believed that participants grasped technical concepts because participants reported that they were doing well. However, when instructors asked specific questions about technical concepts or participant projects, instructors began to understand that participants needed more support. While all instructors themselves learned EMOD through self-led hands-on learning and asking their colleagues questions as needed, participants very much had to adapt to this learning style, and instructors ultimately adapted their approaches as well:

> *Because we all had to learn [EMOD] on our own, [and were] thrown into the deep end, I expected it to be a lot more self-led, you know, ask questions when you need to ask questions. The onus is much more on the participants. I think there’s been a lot of things where we’ve had to handhold quite a bit, and I think that there’s a difference in expectation there…I don’t think it at all is that they’re not motivated, and they’re not interested in their projects, it’s just a very different learning style.* (Instructor A, Endline, Cohort 1)

> *At first I was not l meeting them where they were. I think even the way we designed the tutorials and stuff, we realized [were] not the most accessible. We also talked about communication differences around asking questions and working independently versus feeling more comfortable trying things for the first time together. […] We’ve adjusted and realized, [if the participant’s perspective is] “but I flew all the way here to learn something,” maybe it’s to learn it from us directly. And that’s been easy to do.* (Instructor D, Midline, Cohort 1)

During Cohort 1, instructors adjusted some self-led learning activities to be more collaborative and worked to provide instruction and support in ways that resonated with participants. These adaptations included working on example exercises together during tutorials rather than leaving them as homework and instituting formal office hours for dedicated support time. In Cohort 2, instructors knew that they should be proactive in providing support and that participants would need time to feel comfortable with asking questions and making use of the resources available to them. As FEP progressed, Cohort 2 participants grew more comfortable with seeking support during office hours, learning sessions, and ad-hoc one-on-one sessions. Cohort 2 instructors who had previously been participants better understood when and why participants wanted instructor support:

> *The questions that they ask, sometimes it’s just about clarity. For example, writing the aims page, they have a draft, and then they want you to have a look at it and discuss with them. Sometimes we have the journal club meetings, and they want you to see whether what they want to present is okay or not. Sometimes they get stuck with a [coding] script and then they come to sit down with you to see how you can walk them through resolving it.* (Instructor E, Midline, Cohort 2)

### Recommendations for future FEP implementations

Participant feedback and instructor reflections identified opportunities to improve the sequencing and implementation of program materials. The Cohort 1 curriculum first introduced each component that participants would need to build their projects before showing a full example of how components worked together. Upon reflection, instructors and participants felt that having the big picture sooner would help participants understand why different components were important. This change was implemented for Cohort 2.

> *You’re trying to break it down and say, “We don’t need to do everything all at once because everything all at once is going to be way too much, way too overwhelming, and you’re not going to learn any of it.” But not having the big picture to kind of put it into, I think it is a little challenging. You know, just in the sense that, you can make a demographics file, but, like, what does that actually mean? What does that actually do for this? It’s great if you can add a random intervention, but how does that make sense in the context?* (Instructor A, Midline, Cohort 1)

Participants in Cohort 1 provided recommendations to facilitate preparedness for the content and pace of FEP, such programming boot camps:

> *We have been using MATLAB software for modeling. But here, they use Python and R. I think it would also be a good thing to have that in those introductory courses. And maybe have time, that future participants have time to learn those programs in the first two weeks.* (Participant A, Endline)

> *The other thing that I feel is needed is to have very good basic introductions of the programming languages that they are using. It doesn’t necessarily need to be us who are teaching it. […] In our team, almost all of us use one language to run these models, and another language to do analysis. And often we don’t really talk much about the analysis, which is using R (language). So towards the end, they finally start to realize that, there’s a lot of things that you need to use.* (Instructor B, Endline, Cohort 1)

Instructors built on participant feedback from Cohort 1, meeting notes, and post-FEP discussions to prioritize adaptations for Cohort 2. Additional resources for participants before and during FEP were developed, including a participant handbook with a weekly breakdown of to-do lists, detailed session descriptions and objectives, and an overview of journal club articles. New pre-program resources for Cohort 2 included online courses for Python and R, a coding assignment, and virtual sessions on basics of malaria and epidemiology, a journal club, and an alumni advice session. However, participants were much less engaged virtually, and several did not complete the coding exercise.

Some participants in Cohort 2 felt that in-person training time should primarily focus on working on their EMOD projects, and certain scientific development and technical components could be done remotely prior to the in-person portion:

> *I think we can start some aspects of the program before we come. And when we come here, I think it’s more to perhaps, I would say, because here we have some advantage to access EMOD. So concerning about input preparation, DHS data, about journal club, about each aspect which is surrounding the program, I think we can do it before we come here. And when we come here, it’s just EMOD. We come here, every time we have some activity, we know it is to work on EMOD.* (Participant E, Midline)

Instructors did not share this optimism about the effectiveness of virtual instruction.

Cohort 2 participants felt that they would have benefitted from a mid-program break to rest or make progress on their projects with no distractions:

> *I also think that during the program, it could be good to have a one-week break, for example. And during the program, you don’t work every day – some days you’re only working on your project. But if you need to, you could take a one-week break, a week where you’re not even meant to be working on your project. A complete break.* (Participant F, Endline)

Feedback from Cohorts 1 and 2 helped to inform curriculum ideation and program structure discussions for FEP cohorts in 2024 and 2025, which were led by FEP alumni in their home countries.

### Post FEP

FEP participants remain in contact with the Northwestern University team and continue to use EMOD components in their research and instruction, apart from one participant from Cohort 1 who has been lost to follow-up. One Cohort 1 participant returned as an FEP instructor in Cohort 2, then directed FEP Cohort 3a in their home country. Two participants from Cohort 2 returned as FEP instructors, and another one directed FEP Cohort 3b, which was implemented in their home country in French. One Cohort 2 participant included an EMOD-based objective in their PhD thesis. Another Cohort 2 participant is completing a manuscript based on their FEP research project and recently joined an international grant application as the modeler on the study. Participants who directed FEP in their home countries applied to international funding opportunities to support Cohort 4. The Northwestern University team continued to provide support for high-performance computing and program advising.

## DISCUSSION

This study focused on participant and instructor perspectives of a faculty enrichment program on applied malaria modeling. Despite challenges with language, learning style, new disciplines, and time management, participants’ expectations and program objectives were largely met.

Participants gained technical proficiency in programming, mathematical modeling, and EMOD to the extent that some alumni are training other students and faculty in EMOD. Participants also made gains in their scientific communication and proposal-writing skills.

### Establishing relevance to participant needs and priorities

Findings from this evaluation and other previous research suggest that training programs targeting African scholars and practitioners must have strong alignment with local context and needs, provide sufficient professional development to support participant scholarship, and implement adaptive strategies for evolving contexts [7–9]. Previous studies emphasize the importance of co-creation with in-country stakeholders prior to program implementation to ensure that training programs reflect the needs, priorities, and cultural contexts of prospective participants [4,10–12]. Without this, training programs risk being irrelevant to the needs of potential participants and potentially unsustainable.

FEP was designed to address skills gaps in state-of-the-art models and provide participants the scientific communication skills necessary to effectively engage in scientific collaborations with global modelers and researchers. Prior to Cohort 1, FEP program staff sought input from scholars in sub-Saharan African universities to understand the impact of skills gaps on professional advancement and collaboration with national programs. These discussions also revealed the technical, communication, and professional development skills that aspiring applied modelers felt they could benefit from the most. As such, the initial FEP curriculum was deeply informed by the perspectives of prospective participants.

FEP feedback loops at each stage of data collection allowed program staff to implement responsive and adaptive strategies for supporting participants. Studies about similar training programs suggest that evaluations before, during, and after training programs are critical to understanding the full scope of program impact on participant experiences [8,13,14]. Similar to these studies, post-program data collection was anecdotal, with limited metrics for measuring publications from FEP projects or integration into local health systems. Although anecdotal post-program data captured positive indications of knowledge transfer, partnership opportunities, and pending publications, future training programs should consider robust post-program data collection to effectively measure long-term program impacts.

### Cross-disciplinary learning

Cross-disciplinary learning is a valuable tool for addressing global health challenges [15], and bridging knowledge gaps between theoretical and applied fields of study [16] across global health, medicine, engineering, and social sciences. Because the FEP curriculum was cross-disciplinary but FEP participants varied in disciplinary backgrounds, epidemiological knowledge, and experience with programming, all participants were learning in a different discipline at some point in the program, and some participants had to learn quickly in multiple topics simultaneously. FEP instructors had to quickly adapt to address gaps in pre-program resources and preparation. Cross-disciplinary learning was especially challenging for participants with more mathematical backgrounds. A study of practices across STEM disciplines highlighted differences in pedagogical approaches to models, where similar to FEP findings, those with mathematical backgrounds tend to focus on the theoretical aspects of models compared with those from empirical science backgrounds [17].

STEM education studies discuss how cohort learning and peer feedback can improve cross-disciplinary learning, improve critical thinking skills, and encourage collective problem-solving [18,19]. During FEP, participants’ various backgrounds in the different disciplines relevant to applied malaria modeling allowed them to lean on one another when they encountered challenges. While FEP participants developed individual projects, team projects can also help participants whose experiences are challenged by their disciplinary perspectives [20].

### Supporting the scholarship and agency of African researchers

Global health training programs must ensure that scholars, especially those from low-and middle-income countries (LMICs), are not just recipients of knowledge, but active contributors [10,12]. Some studies suggest that this can be partly addressed by remaining cognizant of the challenges that scholars from LMICs face, including challenges with authorship, research visibility, and funding. In the case of FEP, scientific development and presentation skills were essential to providing participants with the tools to expand access to new funding and research presentation opportunities. Findings from FEP and other studies suggest that multi-pronged training approaches can help researchers negotiate more equitable research partnerships and leverage their technical capabilities more effectively [21–24]. Studies also highlight the importance of scientific development components like those included in FEP in helping African researchers advance their scholarship in the absence of funding and grant-writing training opportunities in their home countries [25,26].

In addition to skills training and scientific development, bidirectional learning is an essential component of ensuring that scholars from LMICs remain active contributors during training programs. One framework for measuring sustainable capacity building between high-income countries (HICs) and LMICs suggests that training programs for research capacity building can only be effective if partners in HICs enter the collaboration with the mindset that learning and mentorship will happen in both directions [7]. Other studies also emphasize the role of bidirectional learning in effective scientific partnerships in ensuring that African researchers maintain agency in research and funding collaborations. As researchers in well-resourced institutions improve their understanding of global research and their own biases, they can be more effective collaborators in their partnerships with researchers outside of the United States [9,22,27]. During FEP, program instructors expanded their capacities in scientific communication, instruction, and cross-cultural understanding in addition to strengthening their own technical mastery of applied modeling. Instructors improved their understanding of the contexts in which many of their collaborators work, gained new perspectives on how to communicate with other malaria researchers, and challenged their assumptions about learning norms across global institutions. Future training programs, especially those implemented by institutions in HICs, must include and evaluate bidirectional learning as a core element of program design.

### Knowledge transfer and future directions

As is recommended for sustainability [9,28], the FEP model intentionally sought to create pathways for long-term knowledge transfer and locally-led training programs. Participants saw opportunities to integrate various elements of FEP into curricula and training programs at their home campuses, including journal clubs, EMOD use, and scientific communication. Scientific development was particularly impactful for Francophone participants, who face additional challenges with navigating publication, scientific collaborations, and research visibility [29–31]. These barriers strongly motivated a Francophone participant from Cohort 2 to implement the FEP model in French at their home institution.

The successful transition of the FEP curriculum to in-country universities addresses the need for greater in-country expertise to facilitate additional training [32]. This change removes barriers to FEP participation related to visas, language of instruction, and geography while ensuring that future curricula directly reflect the evolving needs of the local context. Focus on individual competencies without consideration of health system improvements or integration can hamper training effectiveness and limit participant opportunity to apply skills to real-world situations [7,8]. In-country FEP programs provide an opportunity to collaborate more closely with national programs and integrate health systems thinking into future curricula [8,33].

## CONCLUSIONS

This study contributes to the growing body of literature on the importance of cross-cultural, cross-disciplinary training for strengthening capacity in applied analytics for public health. An intense, focused multi-pronged training program can equip aspiring modelers with the necessary technical skills, soft skills, and domain knowledge that can be further transferred in their home institutions, even as new learning environments challenge training instructors and participants alike. Co-creation with prospective participants can ground program components in the needs and realities of global contexts. Bidirectional learning on the part of program staff and continuous feedback from participants allows programs to respond to emerging needs and implement agile improvement strategies during and after the program.

## Data Availability

Data from this study are not availability due to the risk of identifiability.

## LIST OF ABBREVIATIONS

FEP: Faculty enrichment program
NMP: National Malaria Program
LMIC: Low- and middle-income countries
HIC: High-income countries

## DECLARATIONS

## Consent for publication

Study participants provided consent for their responses to be used in publications.

## Availability of data and materials

To protect the privacy and confidentiality of participants, transcripts are not available due to the small sample size and extent of identifiable information.

## Authors’ contributions

Conceived the study: LO, GAO, AS, TH, RR, MR, KT, IA, BM, JG. Collected data: LO, GAO. Interpreted data: LO, GAO, JG. Wrote first draft of manuscript: LO, GAO, JG.

## Acknowledgements

The authors thank the staff at the Institute for Disease Modeling for their support with the development and maintenance of EMOD, and the many members of the malaria modeling community who have engaged on discussions of capacity development over the years.

